# Artificial Intelligence for Hemodynamic Monitoring with a Wearable Electrocardiogram Monitor

**DOI:** 10.1101/2024.04.01.24304487

**Authors:** Daphne E. Schlesinger, Ridwan Alam, Roey Ringel, Eugene Pomerantsev, Srikanth Devireddy, Pinak Shah, Joseph Garasic, Collin M. Stultz

## Abstract

**Background:** The ability to non-invasively measure left atrial pressure would facilitate the identification of patients at risk of pulmonary congestion and guide proactive heart failure care. Wearable cardiac monitors, which record single-lead electrocardiogram data, provide information that can be leveraged to infer left atrial pressures.

**Methods:** We developed a deep neural network using single-lead electrocardiogram data to determine when the left atrial pressure is elevated. The model was developed and internally evaluated using a cohort of 6739 samples from the Massachusetts General Hospital (MGH) and externally validated on a cohort of 4620 samples from a second institution. We then evaluated model on patch-monitor electrocardiographic data on a small prospective cohort.

**Results:** The model achieves an area under the receiver operating characteristic curve of 0.80 for detecting elevated left atrial pressures on an internal holdout dataset from MGH and 0.76 on an external validation set from a second institution. A further prospective dataset was obtained using single-lead electrocardiogram data with a patch-monitor from patients who underwent right heart catheterization at MGH. Evaluation of the model on this dataset yielded an area under the receiver operating characteristic curve of 0.875 for identifying elevated left atrial pressures for electrocardiogram signals acquired close to the time of the right heart catheterization procedure.

**Conclusions:** These results demonstrate the utility and the potential of ambulatory cardiac hemodynamic monitoring with electrocardiogram patch-monitors.

**Plain Language Summary:** Heart failure is a prevalent disorder that is challenging to manage. Part of what makes heart failure management challenging is that the onset of symptoms can be insidious as there are few robust tools that a clinical can leverage to estimate when a patient is likely to experience an episode of acute heart failure. While elevated intracardiac pressures are a reliable harbinger of worsening heart failure, these pressures are typically measured using an invasive, gold-standard approach, which can only be performed in an inpatient setting. A non-invasive method for detecting higher pressures inside of heart would be especially helpful for identifying worsening heart failure in an expeditious manner in the home environment. For this reason, we developed an artificial intelligence model to detect elevated pressures inside the heart using a non-invasive signal, the electrocardiogram (ECG, or EKG), which can be acquired from a wearable patch monitor device. Our results demonstrate that the model provides a reliable platform for the non-invasive assessment of cardiac pressures using data that can be obtained in the outpatient setting.

## Introduction

Heart Failure (HF) is associated with a range of adverse outcomes, resulting in increased healthcare expenditures, morbidity, and mortality^1,2^. Although progress has been made with respect to the management of patients with HF, significant challenges remain. Over 30 percent of HF patients admitted to the hospital are readmitted within 90 days of discharge. Many of these readmissions are avoidable^3,4^.

Patients who have symptoms of congestive heart failure present with elevated left atrial pressure (LAP), requiring medical therapy to reduce these pressures and relieve pulmonary congestion. The gold standard method for measuring the LAP is right heart catheterization (RHC) – an invasive procedure that requires the placement of a catheter attached to a pressure transducer into the right heart and pulmonary arteries. During RHC, a branch of the pulmonary artery can be “wedged”, forming a static column of blood between the catheter and the left atrium. This measurement is referred to as the mean pulmonary capillary wedge pressure (mPCWP) and is a reliable estimate of the LAP and left ventricular diastolic pressure in most patients^5,6^.

Elevated mPCWP is an independent predictor of adverse outcomes in patients with HF, and lowering the mPCWP is an important intervention that can reduce the probability of readmission^7,8^. Furthermore, several studies suggest that the mPCWP begins to rise before the onset of symptoms in HF patients^9-11^. Hence, the ability to track the mPCWP in the outpatient setting would enable physicians to identify high-risk patients and proactively initiate medical therapy, thereby circumventing hospital admission. However, RHC cannot be routinely performed in the outpatient setting. Non-invasive methods for estimating mPCWP suffer from similar challenges that limit their application in an ambulatory setting. For example, while evidence of elevated mPCWP can be garnered from noninvasive cardiac Doppler ultrasound, a skilled sonographer is needed to obtain the required Doppler profiles^12,13^. In addition, accurate estimation of mPCWP from the physical exam alone is challenging and often unreliable, even when performed by seasoned experts^14^. Lastly, simple models that use clinical demographics, including heart rate parameters, perform poorly with respect to LAP estimation^15^.

Due to these limitations, several devices have been developed to estimate mPCWP at home^9,10,16^. The CardioMEMS HF system, for example, is used to measure the diastolic pressure in the pulmonary artery, a surrogate for the LAP, and its use has been shown to reduce the hospitalization rate of patients with advanced heart failure by 50 percent^10^. Despite its benefits in terms of patient outcomes, using CardioMEMS requires an invasive procedure to implant the device within the main pulmonary artery, entailing some risk. The development of a reliable, easy-to-use non-invasive method for monitoring mPCWP, and cardiac hemodynamics more generally, would transform the management of patients with heart failure.

In recent years, artificial intelligence (AI) has shown promise in healthcare applications, including the analysis of medical time-series data to predict patient outcomes. The use of machine learning algorithms can facilitate remote patient monitoring in a range of cardiovascular diseases, via wearable devices measuring the single-lead electrocardiogram (ECG). Previous applications include the detection of atrial fibrillation^17-19^, cardiac ischemia^20^, long-QT syndrome^21^, and reduced ejection fraction^22^. Many of these studies have focused on ECG signals that are acquired from smartwatch devices, which can yield noisy signals when the subject is not at rest^23^. Wearable patch-monitors, which are applied to the chest wall, provide higher quality data, but the utility of ECG signals arising from these devices for machine learning tasks has not been widely explored.

Previously, we proposed a deep learning model that identifies when the mPCWP is elevated using the 12-lead ECG^15,24^. However, the 12-lead ECG is typically obtained during an office visit or in the inpatient setting, and requires the placement of 10 electrodes on the body. Consequently, it is not routinely used for outpatient monitoring. In this work, we propose a system to estimate mPCWP from Lead I of the ECG, called the Cardiac Hemodynamic AI monitoring System (CHAIS). CHAIS uses a deep neural network to analyze a single-lead ECG signal and infer if the patient’s hemodynamics are abnormal. We demonstrate the system’s ability to predict an elevated mPCWP on retrospective datasets of patients from two large hospitals in Boston, MA. We also prospectively evaluate the model on a cohort of patients who wore a commercially available wearable patch-monitor prior to invasive RHC. Using the ground truth mPCWP data from the procedure, we directly evaluate how well the method performs when using data obtained from a wearable patch-monitor.

## Methods

### Retrospective data acquisition

Retrospective data were collected for patients who underwent cardiac catheterization at two institutions (IRB protocol #2020P000132). At MGH, data from right heart catheterization (RHC) procedures performed between January 2010 and October 2020 were collected and matched to the first 12-lead electrocardiogram from the same calendar day as the procedure (N = 6739). Data from the Brigham and Women’s Hospital (BWH) were collected between June 2009 and January 2019, and likewise matched to a same-day ECG (N = 4620). BWH patients who were also provided care at MGH were excluded from the BWH dataset to guarantee non-overlap between training and external validation datasets. Demographic details for the patient populations are provided in Table 1. Samples were further categorized by indication for catheterization using the procedure described in the Supplementary Methods (see Supplementary Table S1).

**Table 1:** Dataset characteristics (ECG = electrocardiogram, STD = standard deviation, mPCWP = mean capillary wedge pressure)

|  | Development dataset | Internal-holdout dataset | External-validation dataset | ECG patch-monitor dataset |
| --- | --- | --- | --- | --- |
| No. ECG signals | 5390 | 1349 | 4620 | 83 |
| No. patients | 3446 | 858 | 2573 | 83 |
| Age at catheterization (mean ± STD) | 64 ± 15 | 61 ± 16 | 57 ± 15 | 67 ± 13 |
| Percentage of samples from male patients | 63.9 | 65.5 | 64.6 | 67.5 |
| Percent with mPCWP > 18 mmHg | 36.3 | 34.3 | 26.6 | 35.0 |

### Data pre-processing

The ECG acquisition machines used at MGH and BWH acquire the ECG signals at either 500 Hz or 250 Hz; we resample all signals recorded at 250 Hz using linear interpolation to 500 Hz. ECGs containing voltage values greater than 5 mV in magnitude were removed, as these likely represent artifacts. For model training, each ECG lead was standardized (Z-scored) using that lead’s mean and variance.

Our original model focused on predicting several hemodynamic parameters from the 12-lead ECG. In this study, we focus on performance of the model with respect to detecting a mean pulmonary capillary wedge pressure (mPCWP) greater than 18 mmHg.

### Model pre-training

CHAIS is a single-lead adaptation of the 12-lead ECG residual neural network for estimating cardiac hemodynamics.^15^ We initially pre-train the model to regress the PR, QRS and QT intervals and the heart rate from the 12-lead ECG, using a cohort of 242,216 patients at MGH. There is no overlap between the patients in the pre-training cohort and those in the hemodynamics-matched MGH development and internal holdout datasets. More details on the model parameters and the model architecture are provided in the Supplementary Methods and in Supplementary Figure S1.

### Fine-tuning on single-lead data

To fine-tune on single-lead data, a single lead of the ECG is replicated 12 times to form a tensor that is 12 x 5000 samples (10 seconds at 500 Hz). Empirically, this procedure yielded better results relative to using a single 1x5000 tensor as input. The final 12x5000 tensor was Z-scored before training and testing.

For fine-tuning on the retrospective single-lead data, the pre-trained model, up to the 416-node layer, is appended with several dense layers (Figure S1c), which are randomly initialized. Additional training details are provided in the Supplementary Methods.

The data from MGH were divided into a development set and an internal-holdout set, with 80 percent in the former (5390 samples) and 20 percent in the latter (1349 samples). The development set is split into training (80 percent, 4304 samples), validation (10 percent, 546 samples), and internal-test (10 percent, 540 samples) sets. The whole BWH dataset is used solely for validation, and is referred to as the external-validation set (4620samples). There are no overlapping patient data in any of these datasets (development, internal-holdout, and external-validation); i.e., data from a single patient only appears in one dataset.

### Prospective data collection

Prospective data were collected from patients scheduled to undergo cardiac catheterization with IRB approval (IRB protocol #2016P001855). Demographic details of these patients are provided in Table 1, and further categorization by indication for catheterization are presented in Supplementary Table S2. Our prospective study uses a commercially available ECG monitor (QOCA Portable ECG 101 Patch-monitoring Device). The monitor records single lead ECG data in 12 bits resolution at a sampling rate of 256 Hz. Eligible inpatients at the MGH who were admitted to the cardiovascular step-down unit, and who were scheduled for a right heart catheterization, were consented by clinical research staff. The ECG patch-monitor device was placed below the left clavicle approximately 24 hours before and removed the morning of their scheduled procedure. The device was oriented to achieve ECG signals similar to ECG lead I. Once removed, information from the device was downloaded to an encrypted laptop and uploaded to a secure server for analysis.

### Patch-monitor device data pre-processing

ECG signals from the patch-monitor device were resampled to 500 Hz, to match the sampling rate used in the 12-lead ECGs on which the model was originally trained. As CHAIS is trained on 10-second segments of ECG data, our goal was to identify 10 seconds of high-quality signal from the patch-monitor that could be used as input to the CHAIS algorithm. Towards this end, we segmented the ECG data arising from the patch-monitor into 5-minute intervals and identified the 10-second within each interval that had the highest signal to quality index (SQI), which is calculated using the NeuroKit2 Python package^21^. If no 10-second segment within a given 5-minute interval was above 0.5, then that 5-minute interval was discarded. Additional details regarding the pre-processing and quality evaluation of the patch-monitor device data are provided in the Supplementary Methods.

### Zero-shot transfer learning to patch-monitor device data

We applied CHAIS to the patch-monitor device data with no fine-tuning. The patch-monitor device ECGs were pre-processed in the same manner as those for the retrospective studies: each high-quality 10-second ECG segment was normalized by its mean and variance in voltage. Probability values were inferred from the pre-processed samples. We used the highest quality 10-second ECG segment from each five-minute interval in the dataset, wherever a sufficiently high-quality segment was available, yielding approximately one CHAIS prediction for each 5-minute interval.

### Evaluation Metrics

To assess how CHAIS could be used in practice, we computed the sensitivity and specificity using the internal and external datasets. To compute the sensitivity, one must first choose a cutoff for the model output that defines a positive prediction – i.e., when the mPCWP is predicted to be greater than 18 mmHg – and uses this value to compute the true positive rate (sensitivity) and the true negative rate (specificity). These cutoffs were derived from the combined development dataset, then applied to the internal holdout dataset.

PPV and NPV can be computed for different population prevalence, or pre-test probability, values, using the following expressions^25^:

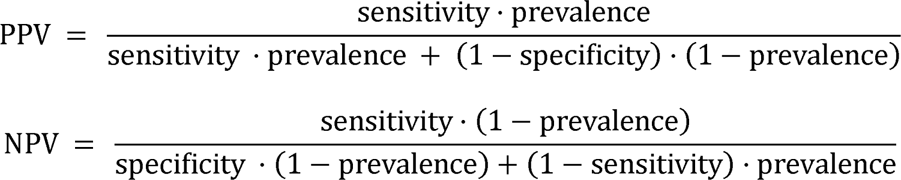

We calculate the area under the receiver operating curve (AUROC) for presenting the model performance on the internal-holdout and the external-validation datasets. For the wearable-ECG dataset from the prospective study, we calculate the AUROC as a function of time to the RHC procedure for respective patients. For each patient, using the method presented above (see subsection *Patch-monitor device data pre-processing*), we acquire one 10-second ECG signal for every 5 minutes over the whole duration of the hospital stay. From the absolute time of the RHC for that patient, we map each of those 10-second ECG to its corresponding relative time to RHC, referred here as the time-delta. This mapping allows us to look back at varying time-deltas from the RHC reference point. We look back at time-deltas ranging from 1 to 24 hours prior to the RHC at an interval of 5 minutes. As shown in Supplementary Figure S4, the number of patients who were monitored at a specific time before their RHC varies with the time-delta, hence the number of available ECG (N) varies with time-delta. For each of those time-deltas to RHC, we then calculate the AUROC over the available ECGs at that time-delta as presented in Figure 4. Only time-deltas for which at least 10 samples were available were included. Error bars were computed as the standard error of the mean over 1000 stratified bootstraps for that data at each time-delta. Standard deviations for AUROC values and other statistics are computed over 1000 bootstraps, where the observed label prevalence is preserved within each bootstrap. Reported error bars in each table and figure correspond to standard error of the mean.

### Model trustworthiness score

To determine when the model is likely to yield a misleading result, we use the Shannon Entropy, in a manner similar to the method used in Raghu et al.^26^ Let f_y_ (x) denote the model probability for one of the inference tasks, y, where x is a given input ECG. Then the entropy for a given prediction is:

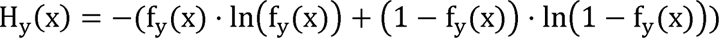

This expression captures, in essence, how close the output model probability is to 0.5, with higher H_Y_ reflecting a value closer to 0.5, and lower model trustworthiness. Note that 0 ≤ H_Y_ ≤ ln 2. The entropy can be used to compute uncertainty for any of the four model targets, independently. To threshold the uncertainty score, we use the value that splits the development dataset into the 90 percent lowest and 10 highest by uncertainty, so as to exclude only the most uncertain predictions.

## Results

CHAIS is trained to detect an elevated mPCWP from single lead ECG data. Four datasets were used to develop and evaluate CHAIS. The development dataset, obtained from Massachusetts General Hospital (MGH), was used to train the model (Figure 1a). An internal-holdout dataset, also obtained from MGH, and an external-validation dataset, obtained from the Brigham and Women’s Hospital (BWH), were used to evaluate the model (Figure 1b). Finally, we prospectively collected single lead ECG data using a commercially available ECG patch-monitor to further evaluate CHAIS performance on this ECG patch-monitor dataset (Figure 1c). Characteristics of patients in each of these datasets are shown in Table 1, and the diagnostic breakdown of each dataset is shown in Supplemental Table S2.

**Figure 1:**
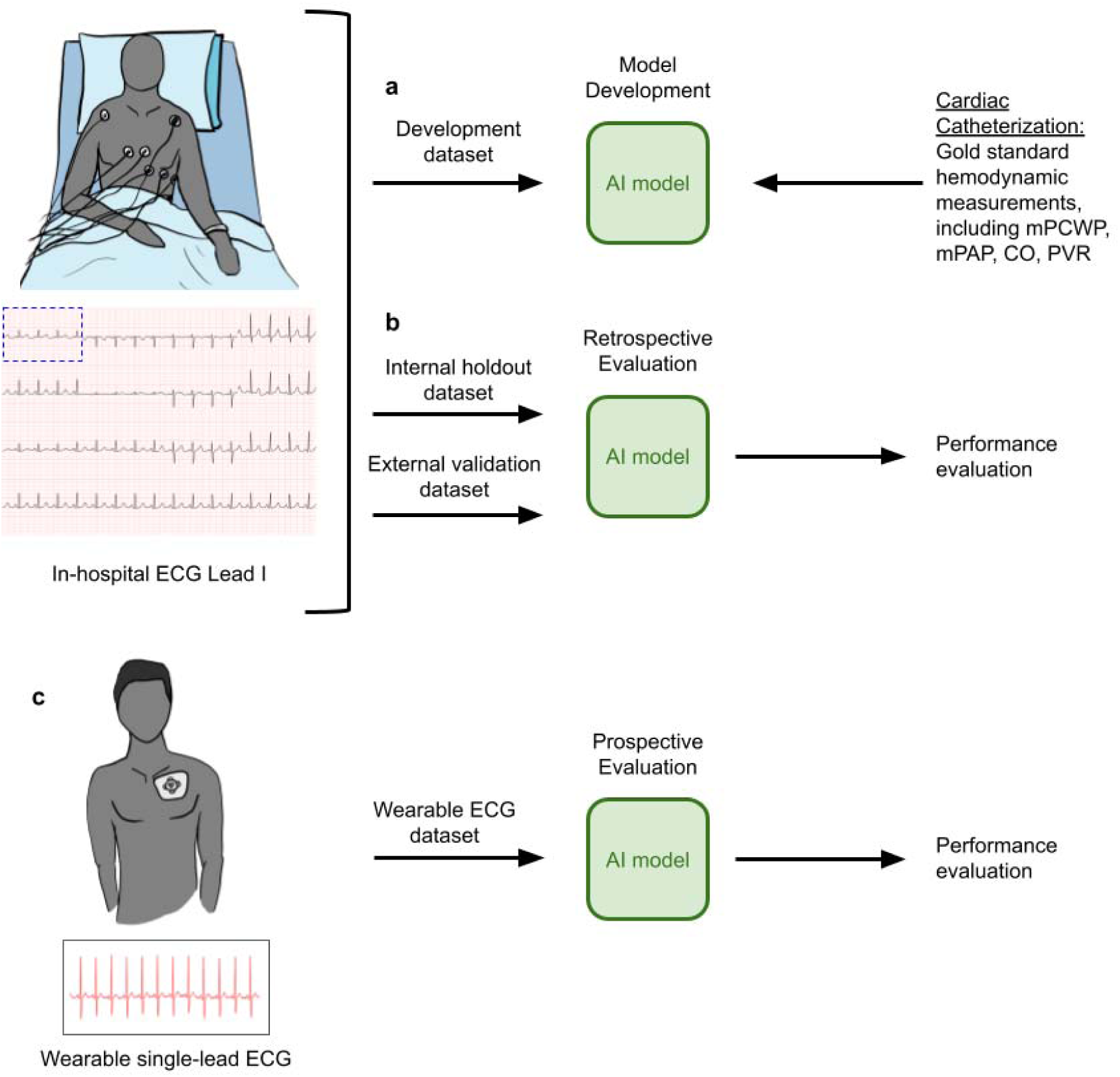
Model development and evaluation. (a) CHAIS is trained on an internal MGH development dataset, using 10s, single lead recordings (Lead I) extracted from the 12-lead electrocardiogram from patients with known cardiac hemodynamic measurements; (b) CHAIS is evaluated on an internal holdout dataset and an external validation dataset containing patients with ECG data (only lead I is used) and known cardiac hemodynamics; (c) the model is then prospectively evaluated using ECG data acquired from patients who wore a commercially available ECG patch-monitor.

### Evaluation on Internal-holdout Set

CHAIS was trained using Lead I of the ECG, as this lead is commonly acquired by patch and wearable ECG monitoring devices^27-30^. We first evaluated CHAIS using an internal-holdout dataset, which does not contain any patients from the development set. For detecting elevated mPCWP, the model achieved an AUROC of 0.80 on the internal-holdout dataset (Table 3).

**Table 2:**
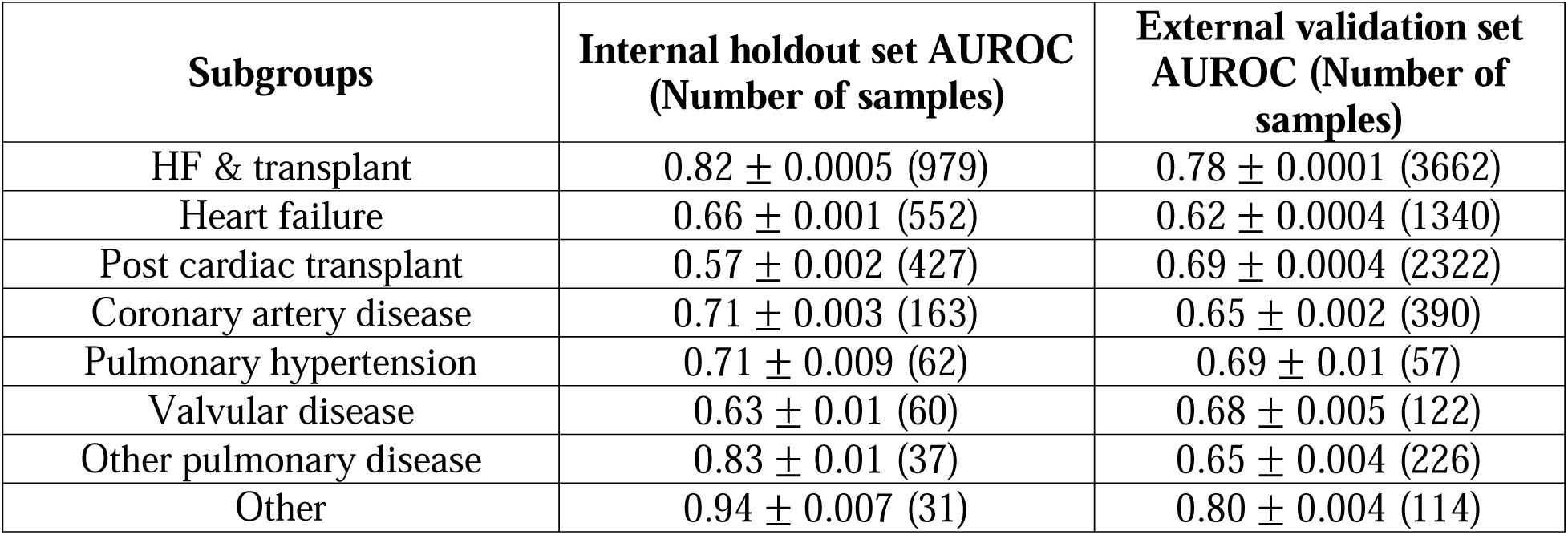
AUROC within subpopulation in the internal holdout dataset for detecting elevated mPCWP. Subgroups with more than 10 samples in both datasets, and at least 1 point in each class, are included (HF=heart failure). Error values correspond to standard error of the mean.

| **Subgroups** | **Internal holdout set AUROC**<br>**(Number of samples)** | **External validation set**<br>**AUROC (Number of**<br>**samples)** |
| --- | --- | --- |
| HF & transplant | $0.82 \pm 0.0005$ (979) | $0.78 \pm 0.0001$ (3662) |
| Heart failure | $0.66 \pm 0.001$ (552) | $0.62 \pm 0.0004$ (1340) |
| Post cardiac transplant | $0.57 \pm 0.002$ (427) | $0.69 \pm 0.0004$ (2322) |
| Coronary artery disease | $0.71 \pm 0.003$ (163) | $0.65 \pm 0.002$ (390) |
| Pulmonary hypertension | $0.71 \pm 0.009$ (62) | $0.69 \pm 0.01$ (57) |
| Valvular disease | $0.63 \pm 0.01$ (60) | $0.68 \pm 0.005$ (122) |
| Other pulmonary disease | $0.83 \pm 0.01$ (37) | $0.65 \pm 0.004$ (226) |
| Other | $0.94 \pm 0.007$ (31) | $0.80 \pm 0.004$ (114) |

**Table 3:**
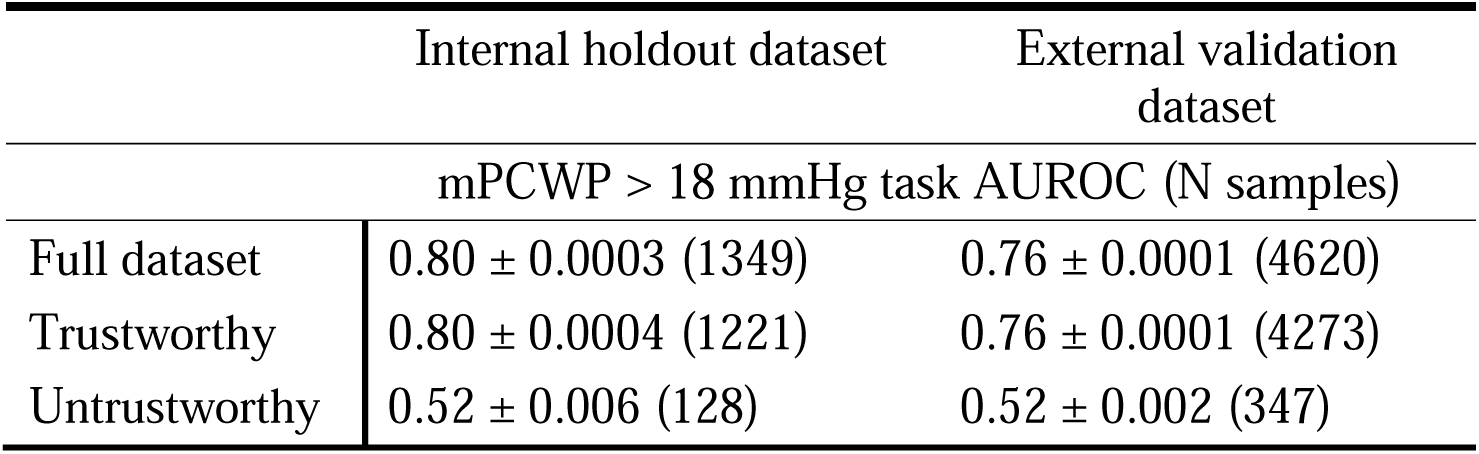
Model performance for detecting an elevated mPCWP ± standard error of the mean. The AUROC are calculated for the full dataset, as well as for the “trustworthy” and “untrustworthy” subsets of the predictions where the “trustworthiness” metric is defined as the Shannon entropy of the corresponding prediction. Standard errors are computed over 1000 stratified bootstraps (mPCWP = mean pulmonary capillary wedge pressure, AUROC = area under receiver operating characteristic).

As no model is perfect, it is important, when possible, to identify predictions that are likely to be incorrect. This can help practitioners determine when to trust a model output. Hence, as a secondary analysis, we calculate the Shannon entropy using the CHAIS output to determine when the model is likely to yield a misleading result, in a manner similar to the method used in Raghu et al.^26^ We hypothesized that predictions associated with high entropy are more untrustworthy relative to predictions with low entropy. We define trustworthy predictions as those that have low entropies, where the entropy threshold is derived from the development dataset, as outlined in the Methods. The derived threshold was 0.6913123. This post-facto analysis of the model predictions can distinguish trustworthy predictions against those that are not sufficiently reliable. We observe, as shown in Table 3, that trustworthy predictions correspond to a subset where the model has higher discriminatory ability relative to untrustworthy predictions. For the task of identifying an elevated mPCWP in the internal-holdout set, the AUROC computed for the more trustworthy predictions was also 0.80 (the same as the performance on the whole internal-holdout set), whereas the AUROC is 0.52 for untrustworthy predictions. Such a trust-score can be helpful for physicians in the decision-making process.

We also evaluated the model’s predictive performance. Using a cutoff that achieves a sensitivity of 70 percent in the development dataset, we find a sensitivity of 71 percent, the associated specificity is 75 percent. The associated confusion matrix is presented in the Supplementary Information in Table S3. With a cutoff that achieves a sensitivity of 80 percent in the development set, the sensitivity is 81 percent, and the specificity is 66 percent in the internal-holdout dataset. The range of calculated specificities and sensitivities is shown in Figure 2a.

**Figure 2:**
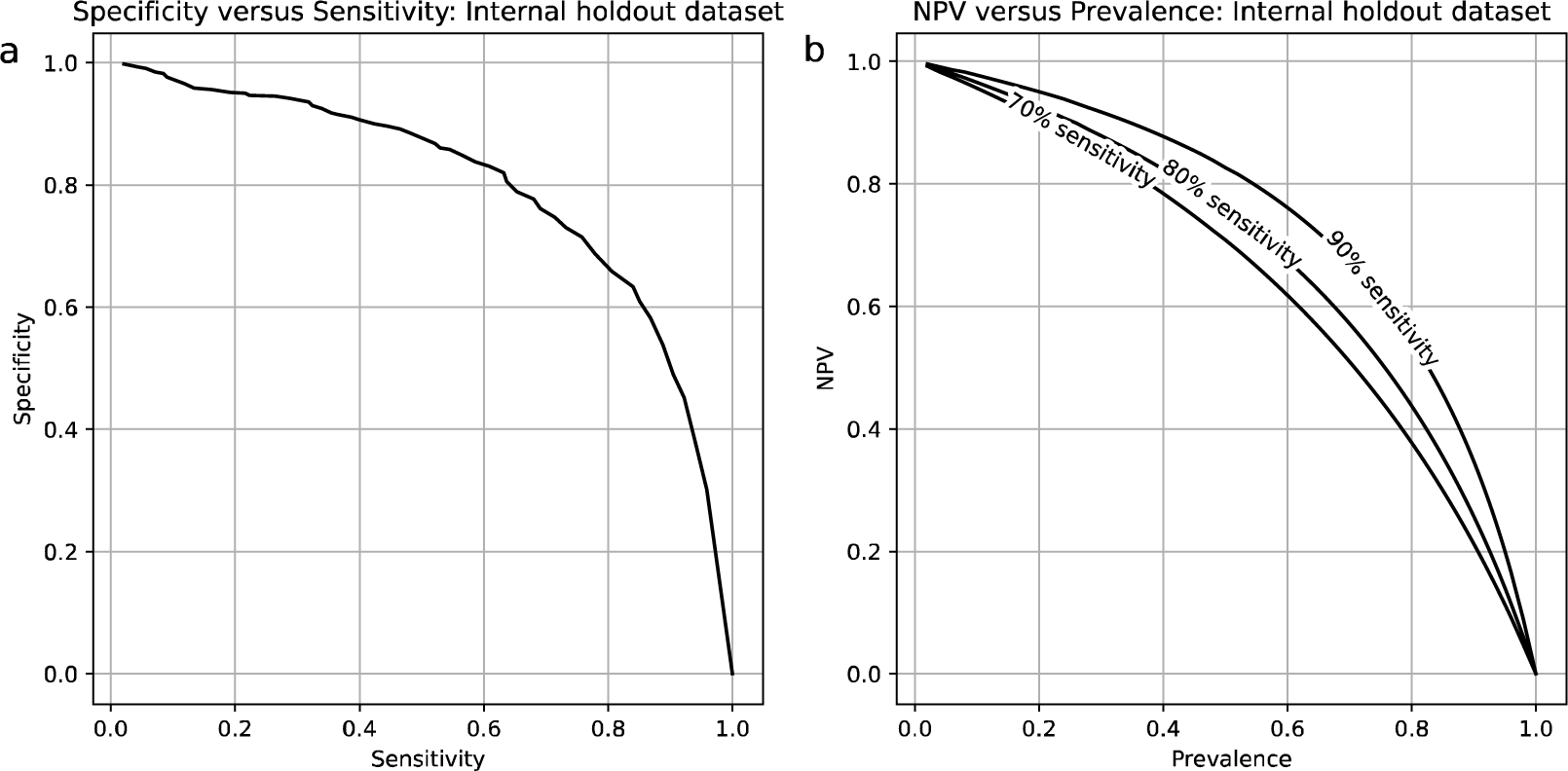
(a) Calculated specificity as a function of the sensitivity using the Internal Holdout Set. (b) Negative Predictive Value (NPV) as a function of the baseline prevalence of elevated wedge in the underlying population (i.e., pre-test probability).

Using these calculated sensitivities and specificities, we computed positive and negative predictive values. As predictive values are a function of the underlying prevalence of an elevated mPCWP in the population of interest, we computed predictive values as a function of different prevalence values. The PPV of the model is generally low and is only above 90 percent when the population prevalence (or pre-test probability) is high; e.g., the PPV is 75.3 percent when the pre-test probability is 50 percent (sensitivity 70 percent).

The NPV as a function of pre-test probability is shown in Figure 2b. When the sensitivity is 80 percent and the pre-test probability is 10 percent, the NPV is 96.6 percent. Yet higher NPVs are attained at higher sensitivities. Exact results for sensitivity thresholds of 70 and 80 percent and for prevalence values of 10 percent, 50 percent, and the observed prevalence are given in the Supplementary Results in Table S4.

### Evaluation on External-validation Set

We further evaluated model performance on an external-validation dataset. For this cohort, the AUROC for detecting a mPCWP > 18mmHg was 0.76 (Table 3). We also calculated the same trustworthiness metric for the model predictions on this dataset for the mPCWP task, as described in previous subsection. The AUROC computed specifically for the more trustworthy predictions was 0.76, and the AUROC computed for the less trustworthy predictions was 0.52 for the task of identifying an elevated mPCWP.

We again computed the sensitivities and specificities for the model for the external validation dataset, using the same decision thresholds as for the internal holdout dataset (Figure 3a). The 70 percent sensitivity threshold (from the development dataset) yields a sensitivity of 55 percent in the external-validation dataset and a specificity of 82.3 percent, while the 80 percent sensitivity threshold (from the development dataset) results in a sensitivity of 68 percent and a specificity of 75 percent. The confusion matrix for the 70 percent sensitivity decision threshold is available in the Supplementary Results (Table S3).

**Figure 3:**
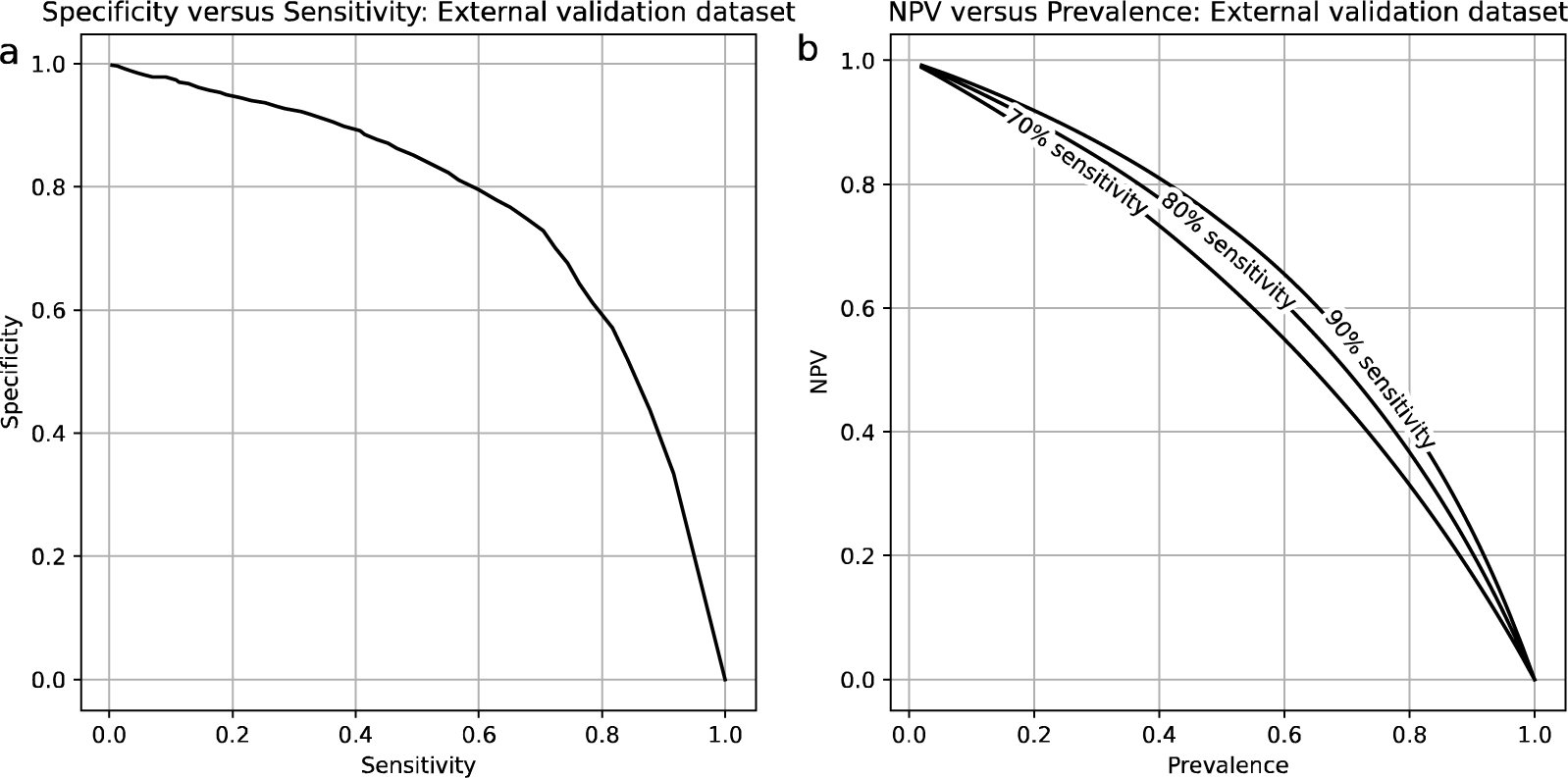
(a) Calculated specificity as a function of the sensitivity using the External Validation Set. (b) Negative Predictive Value (NPV) as a function of the baseline prevalence of elevated wedge in the underlying population (i.e., pre-test probability).

Model performance on the external-validation set in terms of PPV and NPV are given in Supplementary Table S4, for prevalence values of 10 percent and 50 percent and for the observed prevalence in the dataset. The PPV is over 70 percent for a prevalence of 50 percent at sensitivity thresholds of both 70 and 80 percent. The NPV exceeds 90 percent at a prevalence of 10 percent for both sensitivity thresholds reported here. Yet higher NPVs are attained at higher sensitivity thresholds, such as 90 percent. NPV as a function of pre-test probability is shown in Figure 3b.

### Model performance within diagnostic subtypes

We evaluated model performance within diagnostic subgroups. In the HF & Transplant cohort, an important subtype in the context of advances heart failure care, AUROC is slightly higher as compared to the entire dataset. In the internal holdout dataset, the AUROC is 0.82 in this subgroup, and the AUROC was 0.78 in the external validation dataset (Table 2).

### CHAIS performance on ECG patch-monitor data

CHAIS was prospectively evaluated on the ECG patch-monitor dataset obtained from patients who wore a commercially available ECG patch-monitor prior to cardiac catheterization. Here, as before, our goal was to determine if the model can discriminate between patients who had an elevated mPCWP from those who do not.

In this study, the time between the last recorded ECG measurement and the actual time of the catheterization varies from patient to patient because several RHCs occurred later than their scheduled time. Delays in the timing of the RHC happen when other more urgent, unscheduled, catheterizations need to be performed first (e.g. STEMI), or due to a lack of sufficient staffing to perform non-urgent cases. We therefore evaluated model performance relative to the time between the recorded ECG and the time of the actual catheterization. Generally, discriminatory performance increases as the time to catheterization decreases (Figure 4). Evaluating time points where 10 more samples were available, the highest AUROCs are obtained when the ECG data are acquired within 2 hours of the catheterization procedure, with the best AUROC (0.875) at 1 hour and 25 minutes before catheterization. The number of samples available at each time points are shown in Supplementary Figure S4.

**Figure 4:**
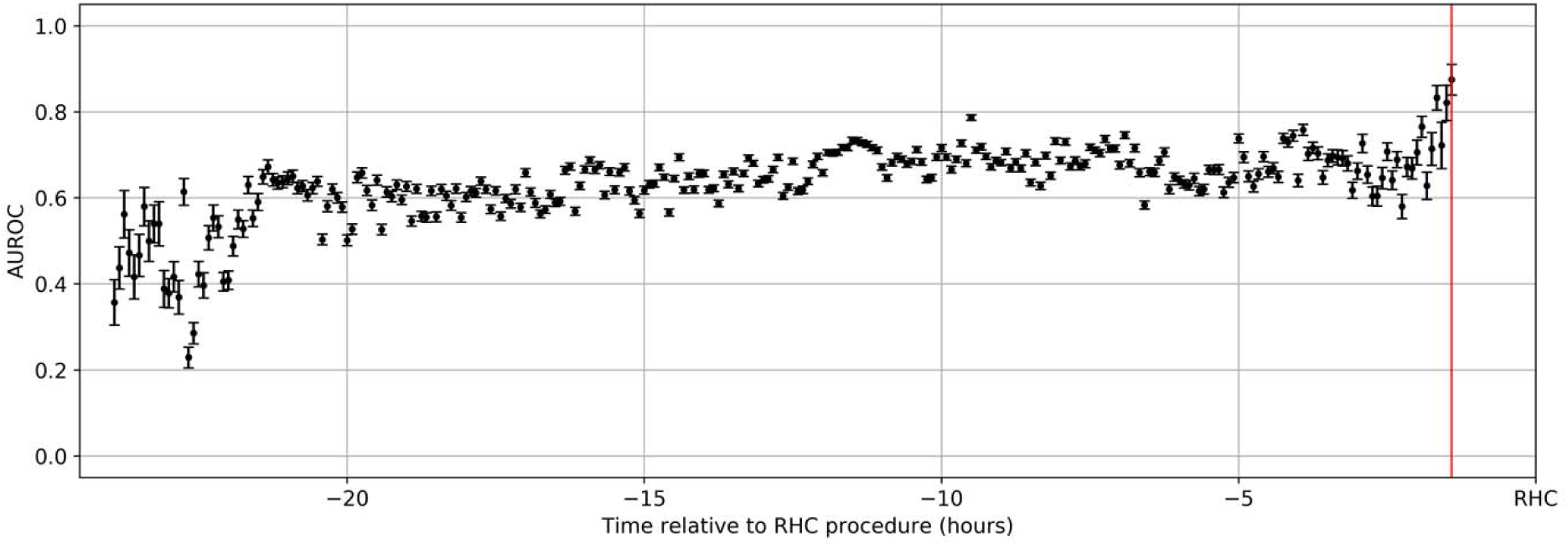
Model performance on the elevated mPCWP detection task versus time relative to catheterization for the ECG patch-monitor data. Samples are extracted from patients where data of sufficient quality is available at a certain time before the catheterization procedure. The plot shows AUROC versus time relative to the catheterization procedure. Error bars correspond to standard error of the mean and are computed over 1000 stratified bootstraps. The vertical line marks 1 hour and 25 minutes before catheterization, the time where the best AUROC is observed. AUCs are reported for time points that have 10 or more observations.

### Intra-patient model performance

We explored the dynamic nature of CHAIS outputs by examining trends in model predictions within the data for individual patients. For the internal holdout dataset, several examples are shown in Supplementary Figure S2. For patients where serial samples were available, predictions appear to track mPCWP on the scale of weeks. In the wearable ECG patch-monitor dataset, we observe significant changes in model outputs on the scale of hours (Supplementary Figure S3).

## Discussion

CHAIS leverages single-lead ECG data to identify patients who have an elevated mPCWP. As the mPCWP rises before the onset of symptoms, non-invasive methods that identify when the mPCWP is elevated enable early identification of patients at risk of developing symptoms of congestive heart failure.

CHAIS was developed and validated using Lead-I ECGs derived from in-hospital 12-lead recordings and tested on prospectively acquired wearable ECG data using a commercially available ECG patch-monitor. CHAIS exhibited good discriminatory performance for detecting elevated mPCWP in the internal holdout (0.80) and external validation datasets (0.76). Calculated predictive values suggest that NPVs arising from the model are informative. The NPV is greater than 95 percent when the pre-test probability is below 10 percent (at a sensitivity of 80 percent or higher) in the external validation set, suggesting that the model can help rule out an elevated mPCWP in low-risk patients.

To determine how the model would perform on data from an ECG patch-monitor device, we prospectively studied patients who wore a commercially available patch-monitor and who underwent RHC approximately 24 hours after monitor placement. CHAIS results on this prospective dataset were obtained in a true zero-shot fashion, with no fine-tuning on the ECG patch-monitor data. Our ultimate goal was to determine if an ECG derived from patch-monitor data can be used to estimate the coincident mPCWP. Consequently, we evaluated whether CHAIS can identify elevated mPCWP when the time between the ECG signal recording and the RHC is minimal, and explore how the discriminatory ability changes as a function of time between the ECG data collection and RHC. We found that the discriminatory ability of CHAIS increased as the time between the ECG recording and the RHC decreased. The AUROC was 0.875 when the time difference between the ECG and the RHC was 1 hour and 25 minutes (the first time where at least 10 data points were available). We were unable to derive statistically robust estimates of the AUROC at shorter time intervals due to the paucity of patients that had ECG at times even closer to RHC. Given the relatively small number of patients in our prospective study, we were also unable to compute reliable sensitivities, specificities and predictive values from these wearable ECG data.

To put the CHAIS AUROC scores into perspective, we note that several studies have estimated the discriminatory ability of echocardiographic measurements for identifying when the mPCWP>18mmHg. In particular, the ratio of the mitral inflow velocity (E wave) and tissue Doppler mitral annular velocity (e’ wave) – quantities frequently measured in echocardiographic studies of patients with heart failure – has been proposed as a robust method for estimating when the mPCWP is elevated^31^. The AUROC of the E/e’ ratio for identifying when the mPCWP>18mmHg has been calculated using small datasets, each containing fewer than 92 patients, yielding AUROCs between 0.68 to 0.78^32-35^. Our method has a discriminatory ability that is on par or better than these estimates and does not require a skilled practitioner to obtain Doppler velocities from cardiac ultrasound.

ECG data from our retrospective cohorts correspond to high-quality Lead I ECG signals. However, the signal quality from wrist-worn ECG devices can be poor relative to the Lead I ECG^23^. By contrast, bipolar single lead wearable ECG patch-monitors, which are applied to the chest wall, typically yield signals comparable to what is obtained with multi-lead high-quality ECG devices^36,37^. Consequently, our prospective study focused on a patch-monitor applied to the chest, rather than a smart watch acquired ECG signal. How these data generalize to signals acquired with wrist-worn monitors remains to be explored. Another limitation of our study is that the prospective evaluation relied on a small cohort of patients admitted for a right heart catherization. Whether these results generalize to ambulatory patients, is therefore not straightforward and requires further prospective validation in larger cohorts. Nevertheless, these results show the feasibility of using patch monitor data for invasive hemodynamic monitoring.

Deep learning models are not inherently interpretable, in part of because of the large number of parameters associated with the models and the nonlinearity of the relationships they are trained to capture. Nevertheless, interpretability is important because it helps users gauge when to trust model predictions, and, in the context of clinical medicine, predictions that are in agreement with one’s prior understanding of pathophysiology are more likely to be trusted. We therefore propose a trust score, which reflects model confidence, and find that more trustworthy predictions are associated with better discriminatory ability for all the tasks we investigate and in all the datasets we examine.

This study describes a deep learning-based method to detect abnormal cardiac hemodynamics from non-invasive ECG patch-monitors. The ability to monitor cardiac hemodynamics in a non-invasive manner would be a transformative addition to the heart failure management toolbox.

## Data availability

Access to de-identified clinical data is only available pending IRB approval.

## Code availability

Model architecture and weights are available online: https://github.com/mit-ccrg/CHAIS

## Acknowledgements

Retrospective data analyses for this study were approved by the Institutional Review Board. (MGH protocol #2020P000132). Prospective data collection and analyses were approved by the IRB at MGH (protocol #2016P001855). We would like to thank Christiana Schneider for her help with data access and study enrollment.

## Author Contributions

DES and RA developed and implemented, and evaluated the described methods. EP, SD, PS, and JG provided data for the study. CMS supervised the work. DES, RA, RR, and CMS contributed to the writing and editing of this article.

