## Supplemental Information for "Artificial Intelligence for Hemodynamic Monitoring with a Wearable Electrocardiogram Monitor"

**Supplementary Methods**

Model Architecture

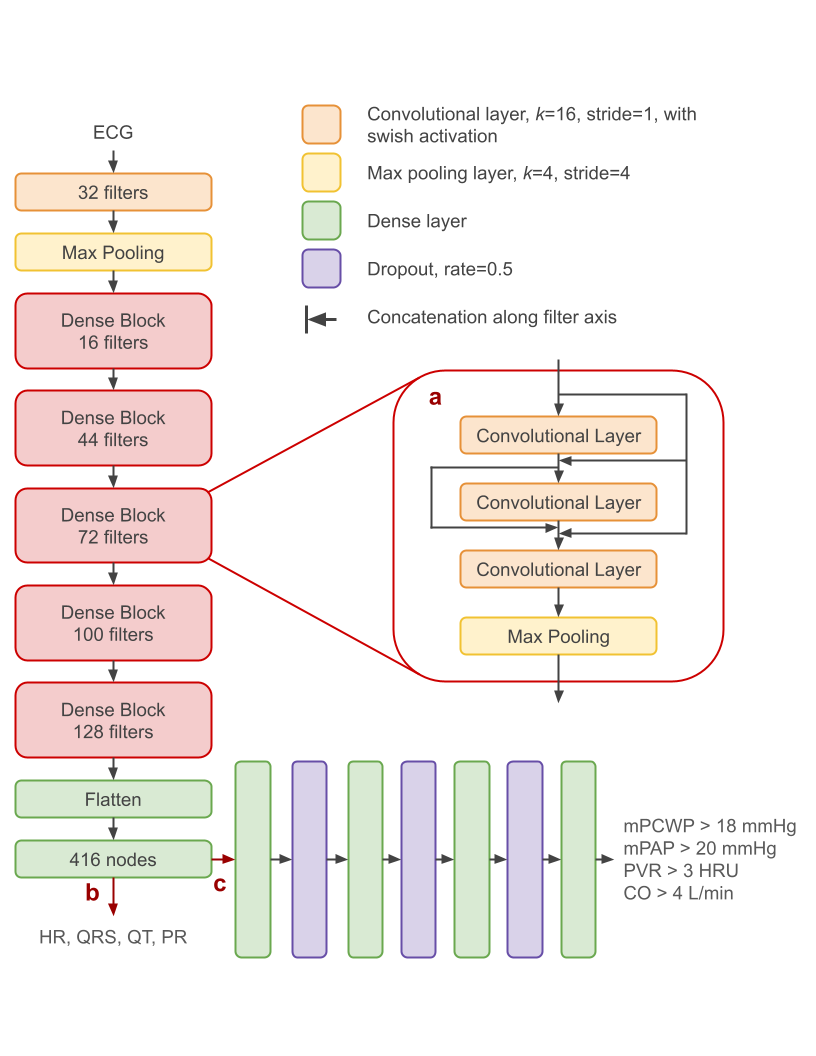

**Hemodynamic predictions**

*Supplementary Figure S1*: Model architecture for pre-training and fine-tuning. The article is a residual neural network, with the characteristic skip connections shown in (a)^1^. (b) During pre-training, the model input is the 12-lead ECG, and the target is to regress intervals (HR and the QRS, QT, and PR intervals). (c) For fine tuning, the model is truncated at the penultimate layer and appended with four dense layers, then trained on single-lead ECGs for the binary tasks listed in the figure.

Training Details

Details of model pre-training are given in Schlesinger *et al*^2^.

The neural network model has 3,042,660 trainable parameters. We trained on the four hemodynamic classification tasks of interest, where the loss function is the sum of the binary cross-entropy (BCE) for each task, using the Adam optimizer^3^. The batch size was 64, and the learning rate was 10^-4^. We trained for 50 epochs, and used the weights from the epoch with the best (lowest) validation loss.

Labeling Diagnostic Sub-Groups

Diagnostic sub-group labels were derived by identifying keywords within descriptive text of a patient’s diagnosis (Table S1). The program checks for keywords without case sensitivity in the order given in the table, and exits (providing a label) once a keyword is detected. A keyword may appear within a longer word, e.g., ‘cardiomyopath’ in cardiomyopathies. For patients with multiple diagnoses, a single label was selected based on the following prioritization: Transplant, heart failure, coronary artery disease, pulmonary hypertension, other pulmonary disease, valvular disease, myocarditis, pericardial disease, peripheral vascular disease, electrical dysfunction, congenital heart disease, and “other.” The method is applicable to the population of patients in this study, namely those that underwent right heart catheterization, as this pre-selects for patients with cardiovascular disease.

*Table S1*: Keywords used to extract general diagnostic labels based on diagnosis descriptive text.

| **Diagnostic Sub-Group Label** | **Keywords** |
| --- | --- |
| Cardiac transplant | ‘transplant’ |
| Heart failure (HF) | ‘failure’, ‘cardiomyopath’, ‘cardiogenic shock’ ‘CHF’ |
| Coronary artery disease (CAD) | ‘coronary’, ‘myocardial infarction’, ‘STEMI’, ‘angina’, ‘CAD’, ‘ischem’ |
| Valvular disease | ‘rheumatic’, ‘valv’, ‘mitral’ |
| Pulmonary hypertension (PH) | ‘pulmonary hypertension’ |
| Other pulmonary disease | ‘pulm’ |
| Congenital heart disease | ‘congenital’, ‘defect’ |
| Electrical dysfunction | ‘rhythm’, ‘fibrillation’, ‘block’, ‘depol’ |
| Myocarditis | ‘myocarditis’ |
| Pericardial disease | ‘tamponade’, ‘pericard’ |
| Peripheral vascular disease | ‘extremities’ |
| Other | (Program reached end of loop without finding any keywords) |

ECG Patch-Monitor Pre-Processing and Signal Quality Index

The QOCA wearable ECG device is worn below the left clavicle of the patient. It records the electrical activity of the heart, namely the Lead-I directional ECG signal. The electrodes of the device capture the differential voltage between the two upper limbs (left arm and right arm) in millivolts (mV) at a sampling rate of 256 Hz. During the study, as the patients wear the device overnight, the signal is continuously recorded with timestamps and is stored on device. The device is removed the following morning prior to the catheterization procedure, and the data was securely transferred to our server for detailed analysis.

The data packets are, first, synchronized with the recorded timestamps and reconstructed as continuous timeseries. To conduct the preprocessing and further analysis, we segment the whole (about 24-hours x 60x60 seconds x 256 samples) timeseries into 5-minutes of non-overlapping windows, each of length 76,800. For each 5-minute segment, we compare the signal amplitude to 5 mV to identify noisy segments and disregard that segment from further analysis. Next, the preprocessing step applies a band-pass Butterworth filter to “clean” signal between 0.05 and 40 Hz. The final step of this data preparation is to calculate the signal quality index (SQI) for that 5-minute segment and identify a best quality 10-second ECG signal.

Using the SQI method implemented in a widely-used ECG analysis Python library NeuroKit2 (<https://github.com/neuropsychology/NeuroKit/>)^4^, we explore the overall quality and the sample-to-sample quality variation of each 5-minute segment. The SQI algorithm segments the QRS complexes from the 5-minute ECG signal. Then, an average QRS complex is computed and the distance of each QRS complex to that average is calculated as a measure of quality. The SQI index is calculated for all samples of the 5-minute segment and represents a relative quality where 1 corresponds to heartbeats that are the closest to the average sample and 0 corresponds to the most distant heartbeat from that average sample. As a limitation of this approach, if the majority of samples are affected by noise or are nonsensical, the average of those cannot represent a good segment leading to all those majority noisy samples getting good SQI scores. In our work, we explored the negative impact of this limitation, and found it relatively insignificant. We apply a threshold of 0.5 SQI index to filter out low-quality samples from 5-minute window. Then, using non-overlapping 10-second windows, we calculate the total SQI over a window, and identify the 10-second window with the best SQI as a representative window from that 5-minute segment. These 10-second ECG windows are then resampled to 500 Hz and used for predicting cardiac hemodynamic abnormalities using the CHAIS model.

**Supplementary Results**

Dataset characteristics

*Supplementary Table S2:* Main indications for catheterization in retrospective datasets. RHC = right heart catheterization

| Main Indication for RHC | Number of cases in Development dataset (percent) | Number of cases in the internal-holdout dataset (percent) | Number of cases in the external-validation dataset (percent) | Number of cases in ECG patch monitor dataset (percent) |
| --- | --- | --- | --- | --- |
| Heart Failure & Transplant | 3980 (73.8) | 979 (72.6) | 3662 (79.3) | 44 (53.0) |
| Heart failure | 2376 (44.1) | 552 (40.9) | 1340 (29.0) | 30 (36.1) |
| Transplant | 1604 (29.8) | 427 (31.7) | 2322 (50.3) | 14 (16.9) |
| Coronary artery disease | 731 (13.6) | 163 (12.1) | 390 (8.4) | 12 (14.5) |
| Valvular disease | 318 (5.9) | 60 (4.4) | 122 (2.6) | 8 (9.6) |
| Pulmonary hypertension | 155 (2.9) | 62 (4.6) | 57 (1.2) | 5 (6.0) |
| Other pulmonary disease | 66 (1.2) | 37 (2.7) | 226 (4.9) | 0 (0.0) |
| Pericardial disease | 20 (0.4) | 0 (0.0) | 22 (0.4) | 1 (1.2) |
| Congenital heart defect | 7 (0.1) | 13 (1.0) | 5 (0.1) | 1 (1.2) |
| Electrical dysfunction | 7 (0.1) | 3 (0.2) | 20 (0.4) | 0 (0.0) |
| Other | 106 (2.0) | 32 (2.4) | 116 (2.5) | 12 (14.5) |

Confusion Matrices

*Supplementary Table S3:* True negative (TN), false positive (FP), false negative (FN), and true positive (TP) counts, derived using a threshold that achieves fixed sensitivity values in the combined internal training and validation datasets.

| Dataset: **Internal-Holdout** N = 1349  **Sensitivity threshold = 70%** Prevalence = 34% | | CHAIS Predictions | |
| --- | --- | --- | --- |
|  |  | Positive  (mPCWP>18) | Negative (mPCWP$\leq$18) |
| Actual Condition from RHC | Positive | 314 | 148 |
|  | Negative | 198 | 689 |

| Dataset: **Internal-Holdout** N = 1349  **Sensitivity threshold = 80%** Prevalence = 34% | | CHAIS Predictions | |
| --- | --- | --- | --- |
|  |  | Positive  (mPCWP>18) | Negative (mPCWP$\leq$18) |
| Actual Condition from RHC | Positive | 360 | 102 |
|  | Negative | 277 | 610 |

| Dataset: **Internal-Holdout** N = 1349  **Sensitivity threshold = 90%** Prevalence = 34% | | CHAIS Predictions | |
| --- | --- | --- | --- |
|  |  | Positive  (mPCWP>18) | Negative (mPCWP$\leq$18) |
| Actual Condition from RHC | Positive | 410 | 52 |
|  | Negative | 410 | 477 |

| Dataset: **External-Validation** N = 4620  **Sensitivity threshold = 70%** Prevalence = 27% | | CHAIS Predictions | |
| --- | --- | --- | --- |
|  |  | Positive  (mPCWP>18) | Negative  (mPCWP$\leq$18) |
| Actual Condition from RHC | Positive | 677 | 553 |
|  | Negative | 600 | 2790 |

| Dataset: **External-Validation** N = 4620  **Sensitivity threshold = 80%** Prevalence = 27% | | CHAIS Predictions | |
| --- | --- | --- | --- |
|  |  | Positive  (mPCWP>18) | Negative  (mPCWP$\leq$18) |
| Actual Condition from RHC | Positive | 834 | 396 |
|  | Negative | 856 | 2534 |

| Dataset: **External-Validation** N = 4620  **Sensitivity threshold = 90%** Prevalence = 27% | | CHAIS Predictions | |
| --- | --- | --- | --- |
|  |  | Positive  (mPCWP>18) | Negative  (mPCWP$\leq$18) |
| Actual Condition from RHC | Positive | 965 | 265 |
|  | Negative | 1317 | 2073 |

*Supplementary Table S4:* Model predictive performance in terms of PPV and NPV at multiple thresholds and with respect to prevalence. (* refers to prevalence observed in dataset)

| **Dataset** | **Sensitivity threshold (percentage)** | **Pre-test probability/ prevalence (percentage)** | **PPV (percentage)** | **NPV (percentage)** |
| --- | --- | --- | --- | --- |
| Internal-holdout | 70 | 10 | 25.3 | 95.6 |
|  | 70 | 50 | 75.3 | 70.8 |
|  | 70 | 34.2* | 61.3 | 82.3 |
|  | 80 | 10 | 21.7 | 96.6 |
|  | 80 | 50 | 71.4 | 75.7 |
|  | 80 | 34.2* | 56.6 | 85.7 |
|  | 90 | 10 | 17.6 | 97.7 |
|  | 90 | 50 | 65.8 | 82.7 |
|  | 90 | 34.2* | 50.0 | 90.2 |
| External-validation | 70 | 10 | 25.7 | 94.3 |
|  | 70 | 50 | 75.7 | 64.7 |
|  | 70 | 26.6* | 53.0 | 83.5 |
|  | 80 | 10 | 23.0 | 95.4 |
|  | 80 | 50 | 72.9 | 69.9 |
|  | 80 | 26.6* | 49.3 | 86.5 |
|  | 90 | 10 | 18.3 | 96.2 |
|  | 90 | 50 | 66.9 | 73.9 |
|  | 90 | 26.6* | 42.3 | 88.7 |

Intra-patient performance

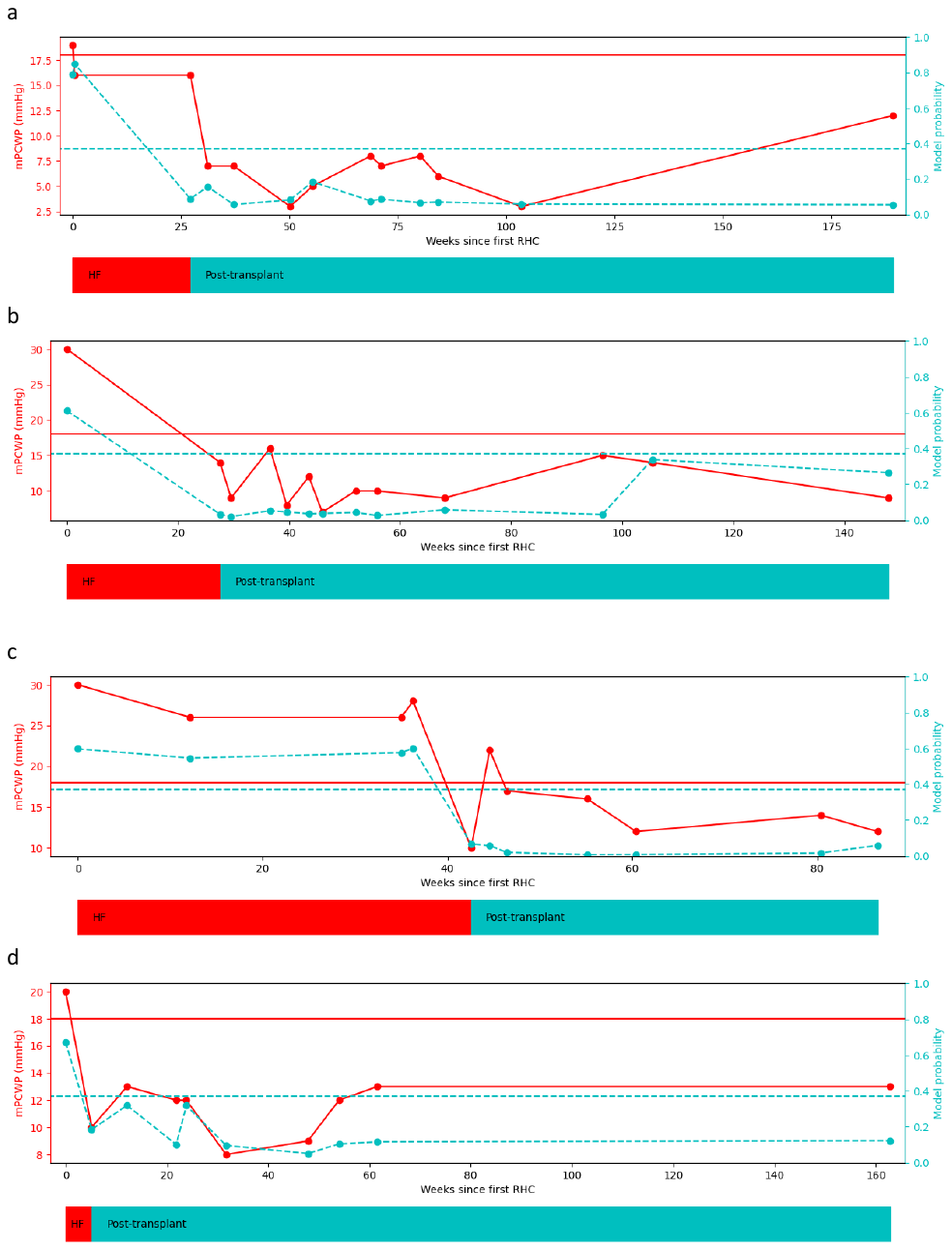

*Supplementary Figure S2:* Examples of trajectories for individual patients from the internal holdout dataset, particularly cases where patients underwent cardiac transplant. The solid, red line with points along it tracks mPCWP on the left-hand vertical axis and the solid, red horizontal line indicates the 18 mmHg threshold. The dashed, cerulean trajectory tracks model output *p*(mPCWP>18 mmHg) on the right-hand vertical axis, and the dashed, cerulean horizontal line marks the probability threshold that achieves a sensitivity of 0.80 in the combined training and internal validation datasets.

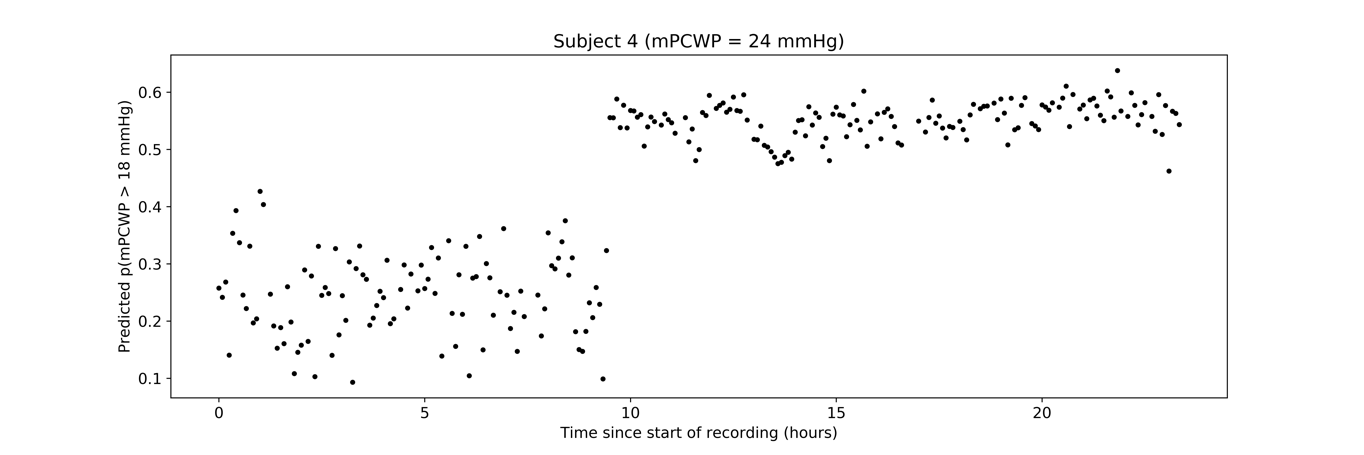

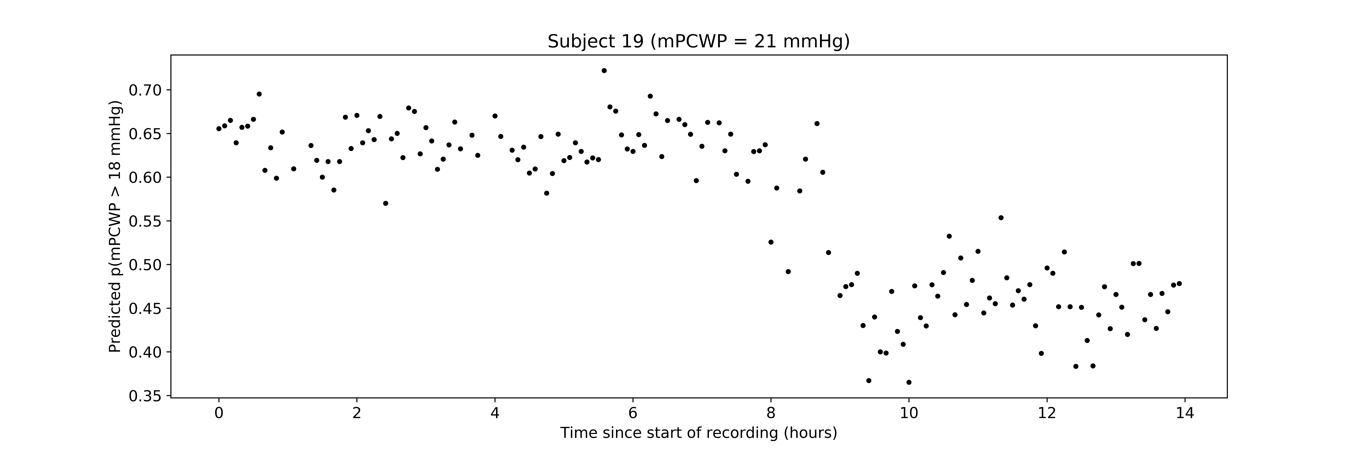

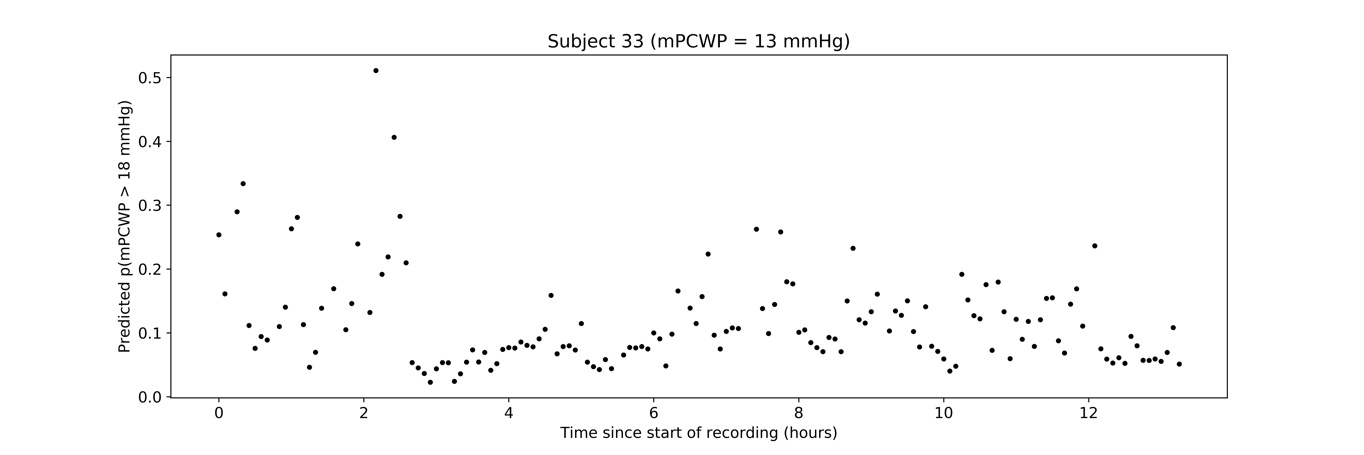

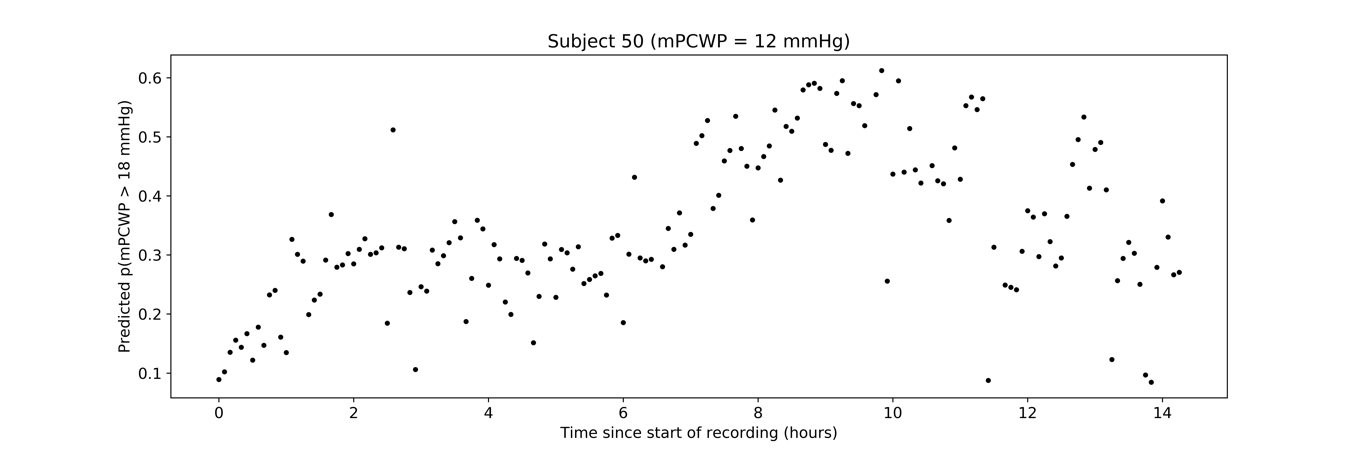

*Supplementary Figure S3:* Intra-patient variability in model predictions for a sampling of patients from the ECG patch-monitor dataset. Subject IDs correspond to de-identified internal ID numbers, which are not known to anyone outside the clinical research staff.

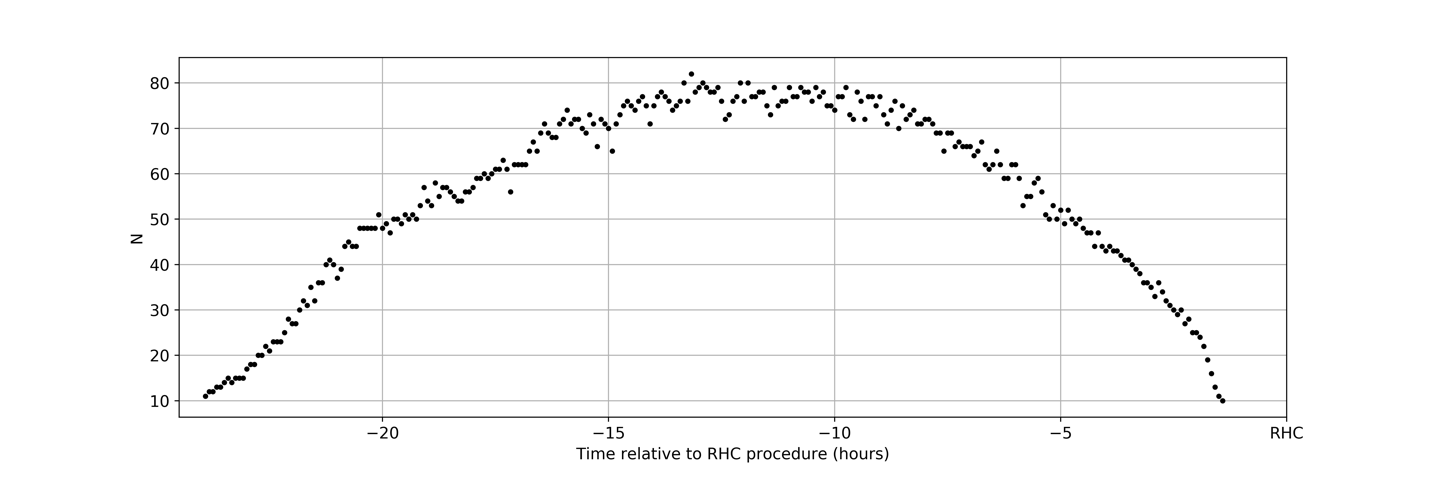

*Supplementary Figure S4*: Number of available samples at each time relative to RHC.
